# Depression risk of 5-alpha reductase inhibitors: A systematic review and meta-analysis with a focus on comparator groups

**DOI:** 10.1101/2025.08.21.25333991

**Authors:** Minh-Ha Nguyen, Tien H Tran, Jeffrey Donovan

**Affiliations:** Department of Epidemiology, Vanderbilt University, USA; Division of Epidemiology, Vanderbilt University Medical Center, USA, Department of Practice, Sciences, and Health Outcomes Research, University of Maryland, USA; Department of Dermatology and Skin Science, University of British Columbia, Vancouver, BC, Canada; Donovan Hair Clinic, Whistler, BC, Canada

## Abstract

**Objective:** To investigate the association of 5-ARI use with depression, specifically focusing on explanations for the heterogeneity in results.

**Design:** Systematic review and meta-analysis

**Data Sources:** A systematic search from Scopus, Embase, and MEDLINE was performed from databased inception to January 2025.

**Eligibility criteria for study selection:** Peer-reviewed empirical studies that reported on measurements of record depression associated with 5-ARIs usage were included. Studies with insufficient data and non-English publications were excluded.

**Data synthesis:** Study characteristics and relative risk estimates were extracted from each article. Estimates were pooled using random-effects meta-analysis. Studies were further stratified by control groups and type of 5-ARI usage to investigate heterogeneity. The Preferred Reporting Items for Systematic Reviews and Meta-analyses (PRISMA) guideline was followed.

**Results:** Five longitudinal studies (n= 2,517,859 patients, effect size = 8), across a wide range of populations (US, UK, Cananda, South Korea) and time-period (1992 – 2018) were included. 5-ARI use was associated with a 31% increased risk of depression (HR 1.31, 95% CI 0.98–1.76), with high heterogeneity (I² = 95.5%, τ^2^ = 0.0984, P < 0.0001). When stratified by comparator type, studies using non-drug controls reported significantly elevated risk (HR 1.61, 95% CI 1.20–2.16, I² = 94.4%, τ^2^ = 0.0635, P < 0.0001), while those using alpha-blocker comparators showed decreased risk (HR 0.89, 95% CI 0.86–0.92, I² = 0%, τ^2^ = 0, P = 0.9711). The choice of comparator group explained the most heterogeneity across studies while the type of 5-ARIs showed similar results (Finasteride HR 1.38, 95% CI 0.51–3.72, I² = 97.4%, τ^2^ = 0.1541, P < 0.0001; Dutasteride HR 1.45, 95% CI 0.51–4.10, I² = 96%, τ^2^ = 0.1688, P < 0.0001).

**Conclusions:** Our findings underscore the importance of comparator selection in pharmacoepidemiologic research and suggest that prior concerns about 5-ARIs may be overstated in studies that use inappropriate control groups.

**Systematic review registration:** PROSPERO CRD42024620917

## INTRODUCTION

5-Alpha reductase inhibitors (5-ARIs), such as finasteride and dutasteride, are commonly prescribed for the management of benign prostatic hyperplasia (BPH) and androgenetic alopecia (AGA). These medications are used by millions of men annually, with finasteride prescribed to approximately 2.6 million men in the United States in 2022^1^. Due to their long-term use in these chronic conditions, there is growing interest in understanding their potential adverse effects, including impacts on mental health.

The proposed mechanism linking 5-ARIs to mood disturbances involves inhibition of the conversion of testosterone to dihydrotestosterone (DHT), which subsequently lowers neurosteroids like allopregnanolone, a GABA-A receptor modulator known to have anxiolytic and mood-stabilizing properties^2^. The literature presents conflicting findings on the relationship between 5-ARIs and depression. Early studies, such as Irwig et al., suggested an association between finasteride use and depressive symptoms post therapy^3^, often termed “post-finasteride syndrome” (PFS), in men treated for AGA. The study was conducted among a subset of 61 men with persistent sexual side effects with self-administrative questionnaire, limiting the generalizability of these findings due to self-selection bias.

Larger cohort studies have since been conducted, producing heterogeneous results^4–8^. Previous meta-analyses^9,10^ reported non-significant pooled results but did not give a sufficient explanation for the observed heterogeneity. To address this gap, we conducted a meta-analysis examining how control group selection contributes to heterogeneity in the reported association 5-ARI use and depression risk. We also provide recommendations for selecting an appropriate control group, depending on study design and research questions.

## METHODS

### Search Strategy and Study Eligibility

The meta-analysis followed a PROSPERO registered protocol (CRD42024620917). Two independent reviewers (MH. N and T.T) systematically evaluated eligibility of all studies retrieved from Scopus, Embase, and MEDLINE for human studies evaluating depression risk associated with 5-ARI usage from inception to January 2025. We screened the reference lists of the articles and where possible, corresponded with study investigators. Full details of the search strategy are provided in the eMethods in the Supplement materials. References identified from database searches were exported to Covidence. Studies presented as abstracts, case reports, conference proceedings, reviews, animal experiments or duplicate publications were excluded. Studies with insufficient data or inadequate reporting and non-English language studies were also excluded. Each full-text article was assessed independently for final inclusion in this systematic review and meta-analysis. Disagreements were resolved by consensus.

### Data Extraction and Quality Assessment

MH. N and T.T independently extracted the following data from each article using a standardized study form: (1) study information – authors, year publication, geographic location, research design, database usage (2) characteristics of participants – mean age, number of exposure, definition of exposure, number of control, definition of control, baseline ascertainment period, mean/median follow-up time, baseline covariates assessment, grace/continuity period, statistical method to control for confounders, and (3) ascertainment of depressive outcomes (number and type of sources of ascertainment). Study quality was assessed using the Newcastle-Ottawa Scale (NOS), with studies scoring ≥7 considered high quality. The rationale for the given scores is on eTable 2 of the supplementary materials. All statistical codes and data will be made publicly available upon publication to promote transparency and reproducibility.

### Statistical Analysis

The primary effect measure was the hazard ratio (HR) with 95% confidence intervals (CIs). Odds ratios were treated as HRs given the low incidence of depression. When multiple HRs were reported due to violation of PH, the larger HR was selected conservatively.

A random-effects model was used to pool effect sizes, with between-study variance (τ²) estimated using the DerSimonian-Laird method^11^. The Hartung-Knapp adjustment^12^ was applied to account for the small number of studies (n=8 effect sizes), improving the robustness of confidence intervals. A random-effects model was selected a priori to account for expected clinical and methodological heterogeneity between studies, including differences in populations, study designs, and outcome definitions. When multiple HRs were reported due to violation of the proportional hazard’s assumption, the larger HR was selected to capture the maximum effect size during initial exposure. Between-study heterogeneity^13^ was assessed using the I² statistic and Cochran’s Q test. I² values of 25%, 50%, and 75% were considered to represent low, moderate, and high heterogeneity, respectively.

Stratification by control groups and 5-ARI agents was conducted to investigate sources of heterogeneity. Stratification by 5-ARI agents was motivated by the difference in molecular weight between dutasteride and finasteride. Dutasteride has larger molecular weight than finasteride 528.539 g/mol^−1^ vs 372.553 g/mol^−1^)^14,15^, which may limit its ability to cross the blood-brain barrier and exert central neurological effects.

We performed leave-one-out meta-analyses to evaluate the influence of individuals. Publication bias was assessed through visual inspection of funnel plots and formally tested using Egger’s test^16^.

All statistical analyses were performed using R version 4.3.0 (R Foundation for Statistical Computing) with the ‘meta’, ‘netmeta’, and ‘metafor’ packages. Statistical significance was set at P < 0.05 (two-sided).

### Public and patient involvement

No patients or members of the public were directly involved in this study as no primary data was collected. Three members of the public were asked to read the manuscript and gave their opinions.

## RESULTS

### Study Characteristics

Five studies involving a total of 2,517,859 patients were included in this systematic review and meta-analysis. These five studies provide eight effect sizes: Garcia-Argibay 2022 (finasteride and dutasteride)^4^, Yeon 2022^7^ (combined 5-ARIs), Hagberg 2017^5^ (finasteride and dutasteride), Welk 2017^8^ (finasteride and dutasteride), and Unger 2016^6^ (finasteride). All were cohort studies, with the exception of Unger et al., which was derived from a preventive randomized controlled trial (RCT) Three of the cohort studies^5–7^ employed a 1–2-year look-back period to minimize prevalent user bias. Most studies included men aged at least 50 years or older, except for Hagberg et al^6^, which included younger participants. The studies were conducted across diverse geographical locations, including Sweden, the United States, the United Kingdom, Canada and South Korea.

### Associations between 5-alpha reductase inhibitors and depressive symptoms

The pooled analysis estimated a 31% increased risk of depression associated with 5-alpha reductase inhibitor usage (HR 1.31, 95% CI 0.98 – 1.76). However, this overall estimate was accompanied by substantial heterogeneity across studies (I² = 95.5%, τ^2^ = 0.0984, P < 0.0001), indicating considerable variability in treatment effects. There were no significant differences in depression risk between finasteride and dutasteride users. Finasteride HR 1.38, 95% CI 0.51– 3.72, I² = 97.4%, τ^2^ = 0.1541, P < 0.0001 vs Dutasteride HR 1.45, 95% CI 0.51–4.10, I² = 96%, τ = 0.1688, P < 0.0001

The leave-one-out sensitivity analysis provided pooled HR ranging from 1.23 to 1.36 with 95% confidence intervals (CIs) spanning from 0.90 to 1.90. These remained stable and consistent with the original pooled HR of 1.31 (95% CI 0.98 – 1.76), indicating no significant influence from any single study. The persistent high heterogeneity (I² 94.4% to 96.05%, τ² 0.0809 to 0.1164) aligns with the original analysis (I² = 95.5%, τ² = 0.0984, P < 0.0001), suggesting that variability is not driven by an outlier but rather by study-level differences.

### Associations stratified by control groups

When stratified by control group, studies comparing 5-alpha reductase inhibitors (5-ARIs) to non-drug users showed a significantly increased risk of depression (HR = 1.61, 95% CI: 1.20– 2.16, I² = 94.4%, τ^2^ = 0.0635, P < 0.0001). In contrast, comparisons to an alpha-blocker/tamsulosin (AB) indicated a decreased risk (HR = 0.89, 95% CI 0.86 – 0.92) with minimal indication for heterogeneity across studies (I^2^ = 0%, τ^2^ = 0, p = 0.9711).

### Publication bias

A funnel plot was generated to assess publication bias (Figure 5). Visual inspection revealed potential asymmetry, with more studies reporting hazard ratios greater than 1 (suggesting increased depression risk with 5-ARIs) and fewer studies with hazard ratios less than 1, particularly among those with smaller standard errors. Egger’s test for small-study effects could not be reliably performed due to the small number of effect sizes (k=8), as the test requires a minimum of 10 studies for adequate power. The observed asymmetry may reflect underlying heterogeneity rather than true publication bias. Nevertheless, this pattern suggests a possible tilt in the evidence toward mild or null effect, as studies with negative results may be underrepresented in the published literature.

## DISCUSSION

This systematic review and meta-analysis of 5 longitudinal studies involving 2,517,859 patients demonstrated that 5-ARI users had a significantly increased risk of depression compared to non-drug users (HR = 1.61, 95% CI, 1.20 – 2.16) while a decreased risk was observed when compared to alpha blocker users (HR = 0.89, 95% CI, 0.86 – 0.92). In addition, when using alpha blocker users as a control group, the pooled HR had little indication for heterogeneity (I^2^ = 0%, τ^2^ = 0, p = 0.9711). Given that most participants in this meta-analysis were aged 50 or older and diagnosed with BPH, alpha-blocker users represent a more appropriate control group than non-drug users, as they better account for the underlying disease burden and age-related risk factors.

When investigating side effects related to a drug that is used to treat an active disease, in this case 5-ARIs for active BPH treatment-it is more appropriate to compare against patients receiving other treatments for the same condition (i.e. active comparator^17^). Making comparison against people with the disease but not taking the medication opens the door for biases by disease burden/indication. As demonstrated by the supplementary data from the appendix of Garcia-Argibay et al^4^, when the risks of depression were lower/non-significant when comparing 5-ARI usage against record AB usage (Finasteride HR 0.82 (95% CI, 0.73 – 0.91) and Dutasteride HR 0.86 (0.70 – 1.02), while the main analysis pointed toward an increased risk of depression associated with record 5-ARIs when comparing to non-drug users (Finasteride HR 1.61, 95% CI 1.47 – 1.76 and Dutasteride HR 1.68, 95% CI 1.43 – 1.97).

This consistency across studies suggests that 5-ARIs are likely not substantially increase depression risk when accounting for underlying disease burden. The use of an active-comparator design helps mitigate confounding by indication, as both groups share similar treatment indications (e.g., BPH), thereby reducing bias from unmeasured confounders such as disease severity or treatment-seeking behavior.

Our study further showed that even with propensity score matching, findings of non-drug control groups are likely subject to confounding by indication. Previous methodological studies have shown that observational studies can replicate placebo-controlled results in preventive trials involving healthy populations^18,19^. Notably, the Unger 2016 study^8^, which reported the smallest increase in depression risk (HR = 1.10, 95% CI: 1.01–1.20), was derived from a preventative RCT evaluating finasteride for prostate cancer prevention over a 7-year period. The modest effect observed in Unger et al., compared to larger effect estimates from observational studies using non-drug users control groups (e.g., Welk 2017 et al, HR = 2.00–2.20; Garcia-Argibay 2022 et al, HR = 1.61–1.68) suggests that even well-matched observational designs may overestimate the association between 5-ARI use and depression due to residual confounding related to underlying disease burden.

The substantial disease burden associated with BPH was highlighted in the cross-sectional study by Pietrzyk et al^20^, which documented a wide spectrum of BPH-related morbidity. We did not include estimates from Pietrzyk et al in our analysis due to challenges in interpretation and concerns regarding methodological limitations. Specifically, the use of stepwise selection procedures for covariates may have inflated the risk of Type I error and compromised the validity of statistical inferences.

## CONCLUSION

Taken together, these findings indicate that while 5-ARIs may exert some biological effect on mood—potentially through mechanisms like neurosteroid modulation—the impact on depression incidence appears to be much smaller than previously reported.

The risk of depression with 5-ARIs use for BPH in men aged 50 and older is likely modest – approximately a 10% increase in risk. The heterogeneity observed in the literature likely stems from differences in control group selection, which may introduce confounding bias due to underlying disease burden, even when propensity score matching is applied. Future studies investigating depression risk in the AGA population should prioritize active comparator designs to mitigate confounding by indication.

**Table 1:**
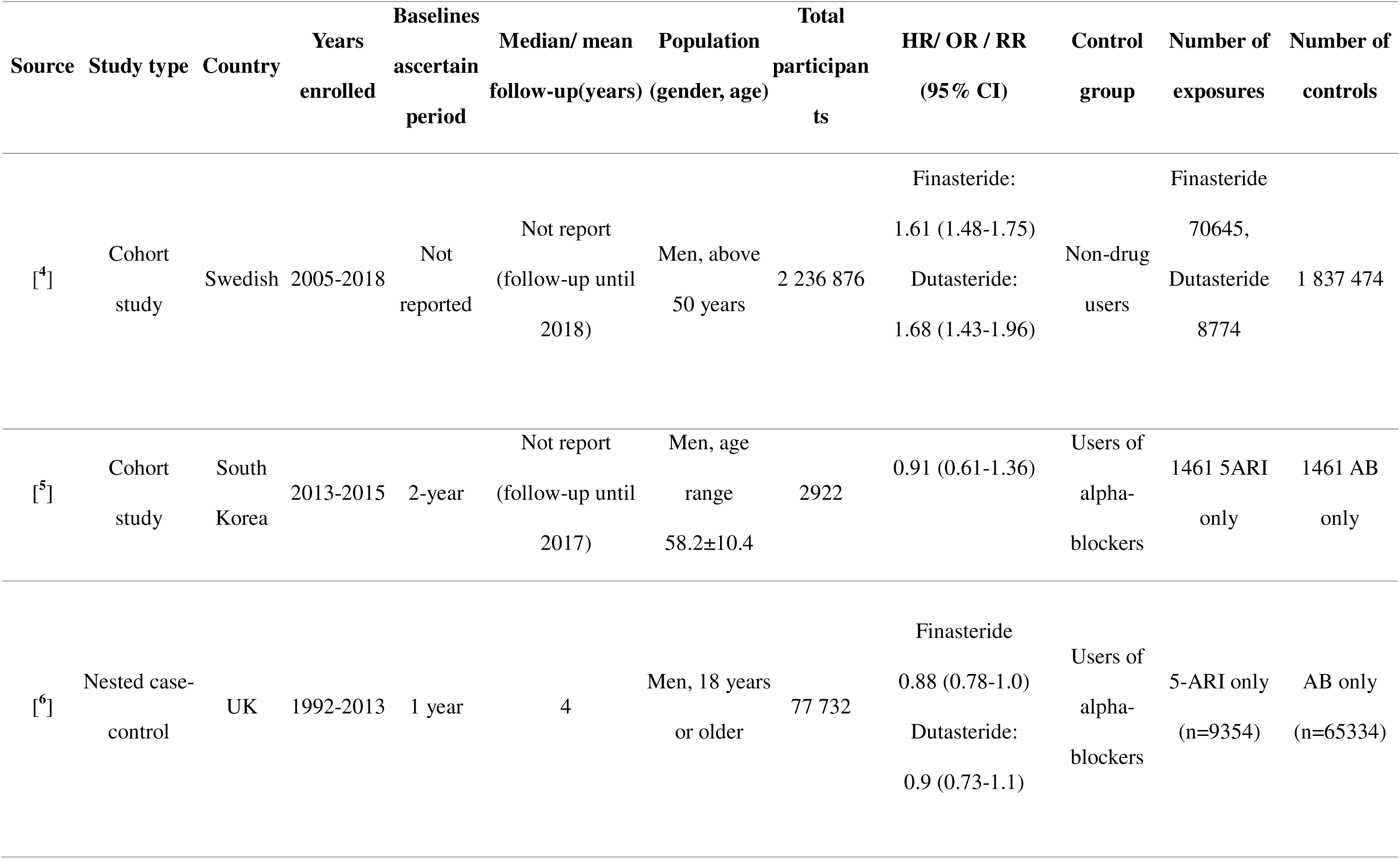
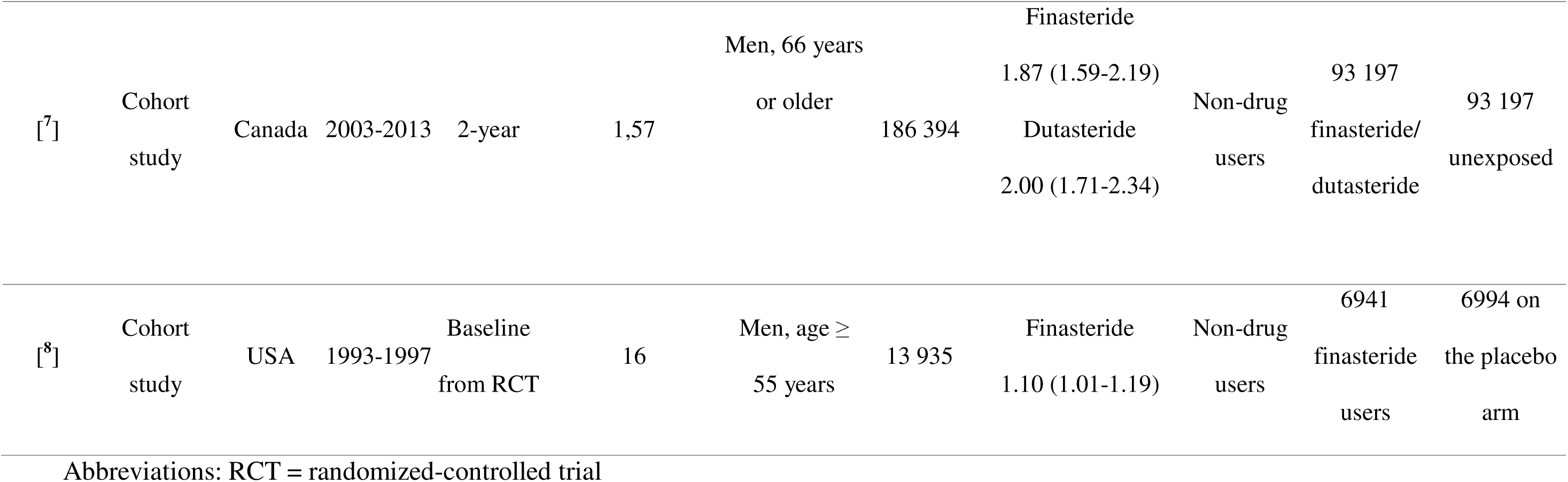
Selected characteristics of the 5 included studies.

**Table 2:** Newcastle-Ottawa Scale score of 5 selected studies.

|  | **Criteria/ Author** | Garcia-Argibay<br>et al. (2022) | Yeon et al.<br>(2021) | Welk et al.<br>(2017) | Hagberg et<br>al. (2017) | Unger et al.<br>(2016) |
| --- | --- | --- | --- | --- | --- | --- |
| Selection:<br>(Maximum 4 stars) | (1) Representativeness of the<br>exposed cohort | * | * | * | * | * |
|  | (2) Selection of the non-exposed<br>cohort | * | * | * | * | * |
|  | (3) Ascertainment of exposure | * | * | * | * | * |
|  | (4) Demonstration that outcome of<br>interest was not present at start of<br>study |  | * | * | * |  |
| Comparability<br>(Maximum 2 stars) | (5) Comparability of cohorts on the<br>basis of the design or analysis | ** | ** | ** | ** | ** |
| Outcome<br>(Maximum 3 stars) | (6) Assessment of outcome |  | * | * | * | * |
|  | (7) Was follow-up long enough for<br>outcomes to occur | * | * | * | * | * |
|  | (8) Adequacy of follow up of<br>cohorts | * | * | * | * | * |
|  | Total score | 7 | 9 | 9 | 9 | 8 |

## Data Availability

All data produced in the present work are contained in the manuscript

## AUTHOR CONTRIBUTIONS

**MH.H:** study conception, literature review, data analysis, writing the first draft and the guarantor of the manuscript

**T.H.T**: study conception, literature review, manuscript review

**J.D**: study design, manuscript review

The corresponding author attests that all listed authors meet authorship criteria and that no others meeting the criteria have been omitted

## Funding

Nothing to disclose

## Role of the Funder/Sponsor

The funders had no role in the design and conduct of the study; collection, management, analysis, and interpretation of the data; preparation, review, or approval of the manuscript; and decision to submit the manuscript for publication

## Disclaimer

The opinions, results and conclusions herein are those of the authors and do not reflect the official policy or position of the funding sources

---

**Figure 1:**
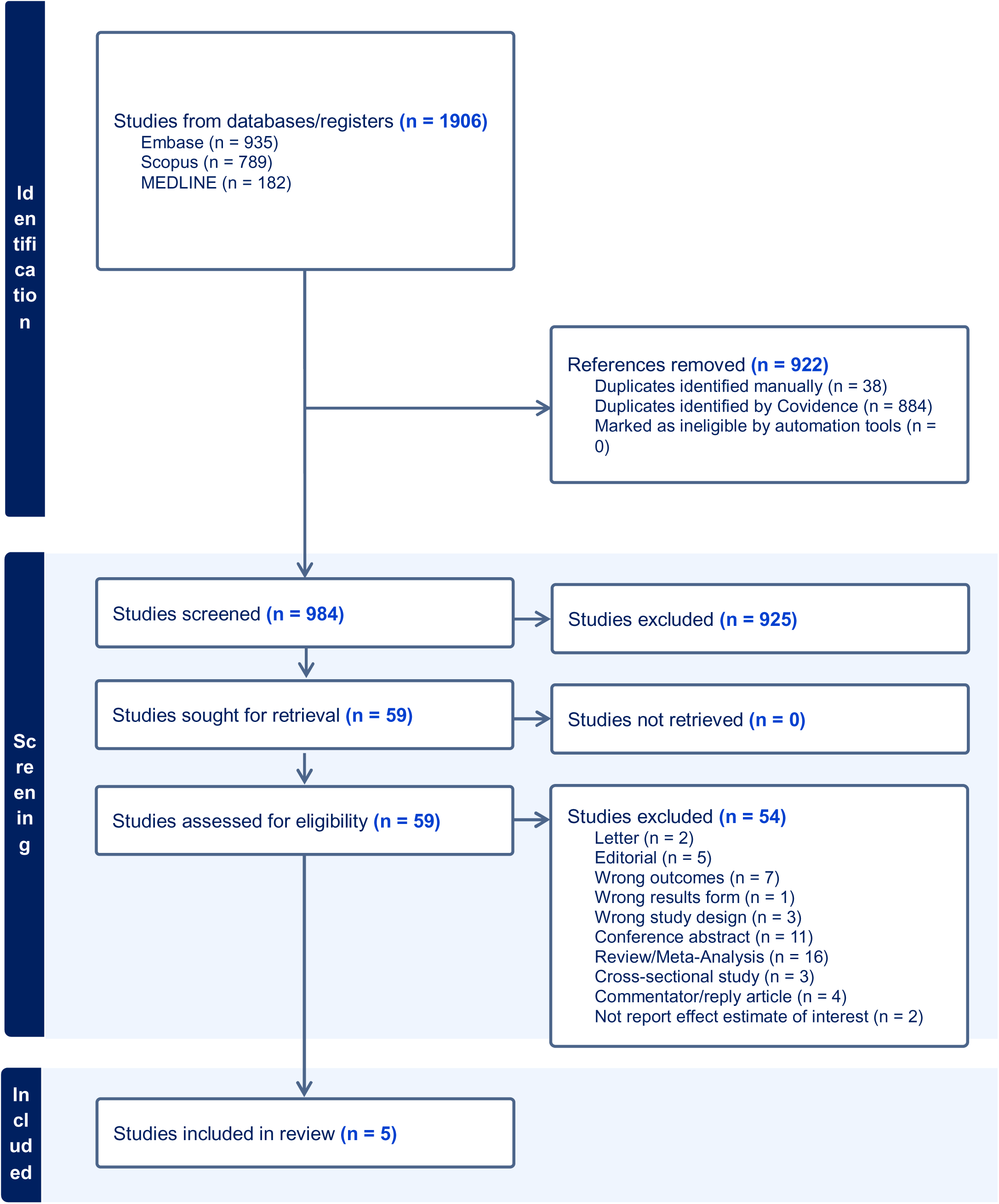
PRISMA Flow Diagram

**Figure 2:**
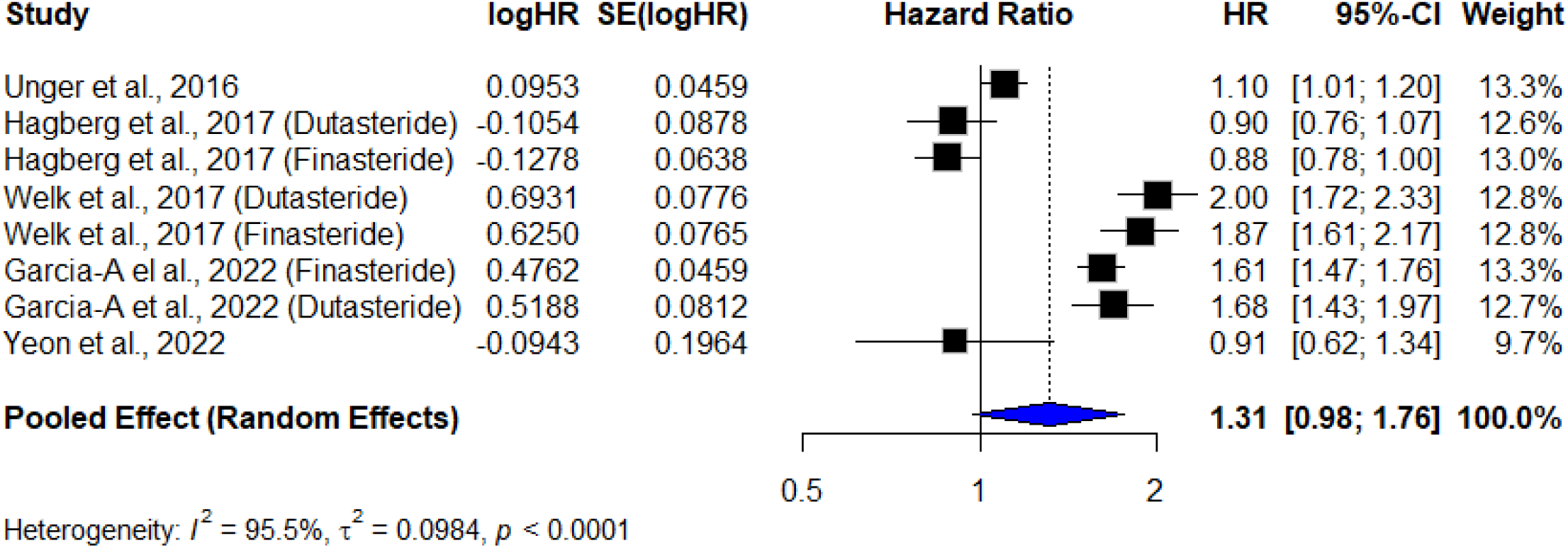
Pooled effect estimates

**Figure 3:**
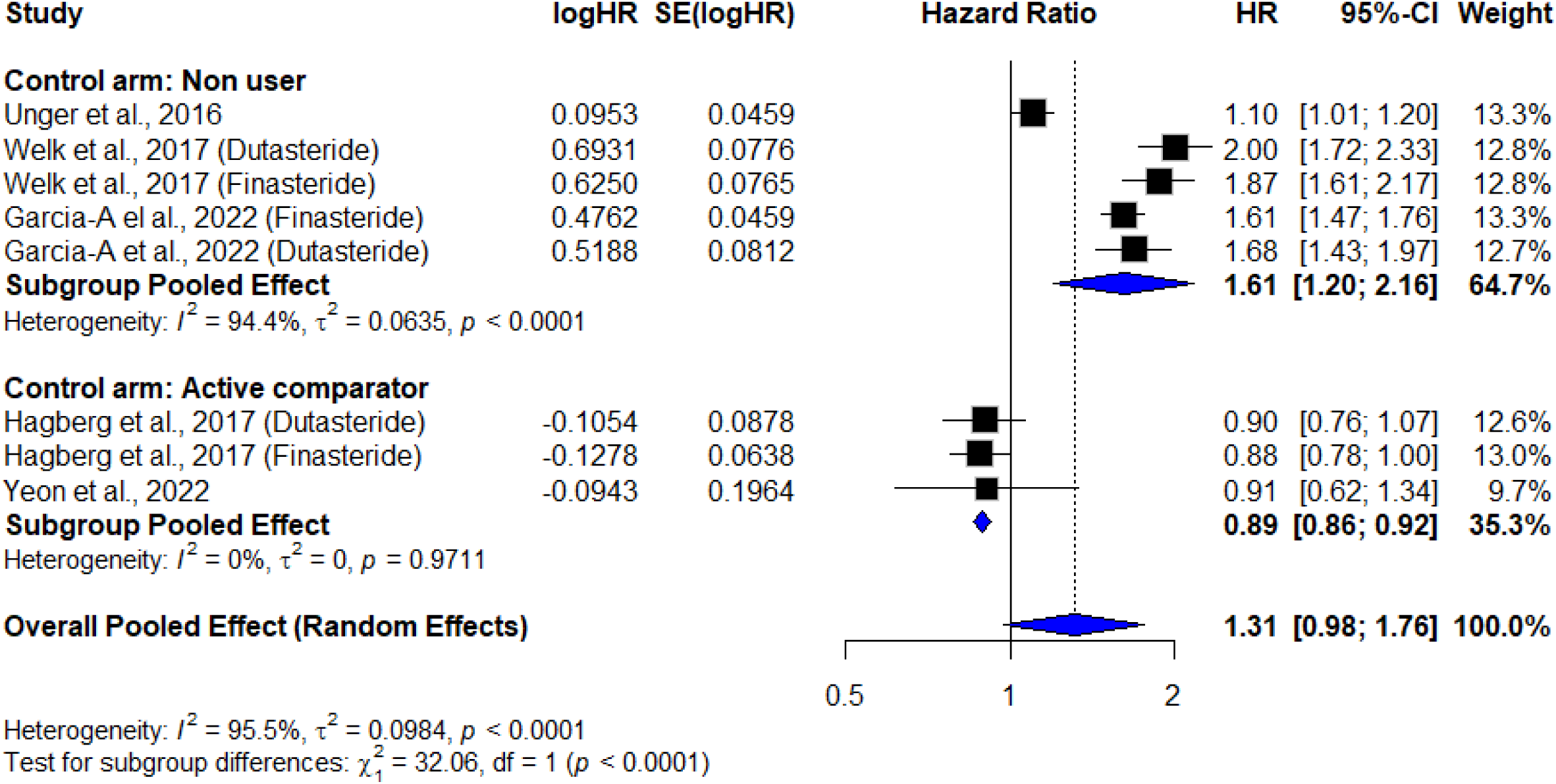
Pooled effect estimates stratified by control groups

**Figure 4:**
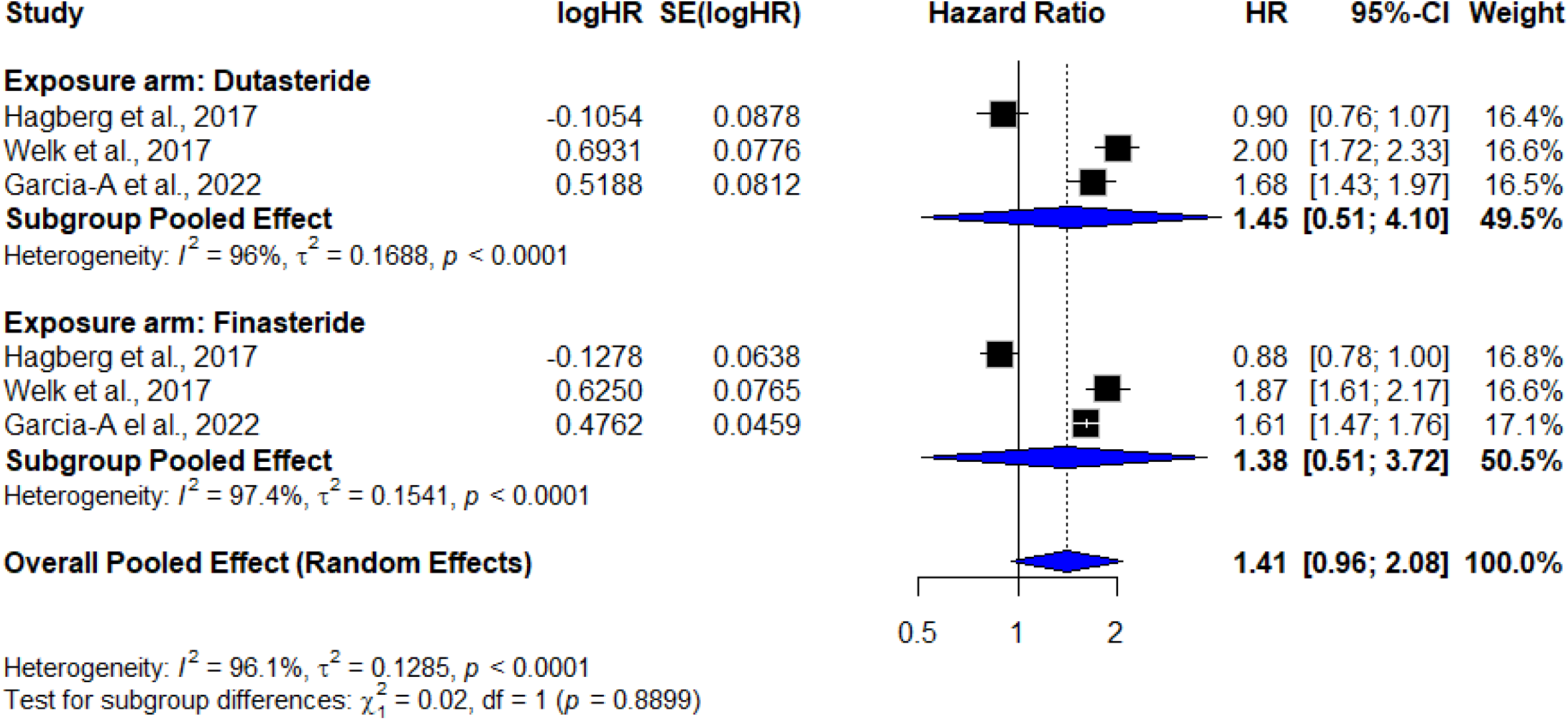
Pooled estimates stratified by 5-alpha reductase inhibitor agents

**Figure 5:**
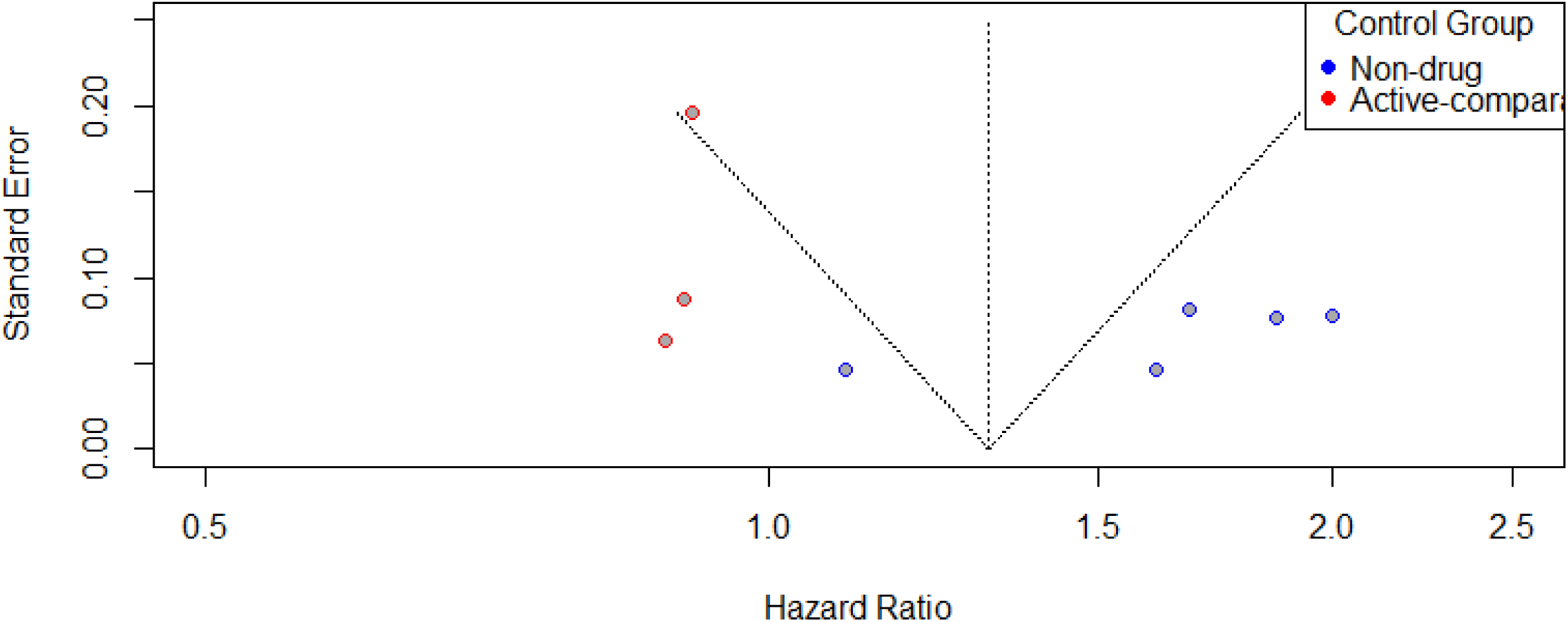
Funnel plot to assess publication bias

## Notes

### Competing Interest Statement

The authors have declared no competing interest.

### Funding Statement

This study did not receive any funding

