## Supplementary materials for "Depression risk of 5-alpha reductase inhibitors: A systematic review and meta-analysis with a focus on comparator groups"

**SUPPLEMENTARY MATERIAL**

This supplementary material contains 9 pages and 2 tables.

**eResults**

eTable 1: Leave one out analysis

| **Results removed** | **Pooled HR** | **CI Lower** | **CI Upper** | **I^2^ (%)** | **Tau^2^** | **Q-value** |
| --- | --- | --- | --- | --- | --- | --- |
| Gracia et al, 2022 (Finasteride) | 1.269 | 0.902 | 1.785 | 95.46 | 0.1164 | < 0.001 |
| Gracia et al, 2022 (Dutasteride | 1.264 | 0.903 | 1.769 | 95.9 | 0.1048 | < 0.001 |
| Yeon et al, 2022 | 1.364 | 0.989 | 1.880 | 96.05 | 0.0986 | < 0.001 |
| Hagberg et al., 2017 (Finasteride) | 1.395 | 1.024 | 1.900 | 94.4 | 0.0809 | < 0.001 |
| Hagberg et al.,  2017 (Dutasteride) | 1.344 | 0.953 | 1.898 | 95.53 | 0.094 | < 0.001 |
| Welk et al., 2017 (Finasteride) | 1.245 | 0.904 | 1.714 | 95.51 | 0.0967 | < 0.001 |
| Welk et al., 2017, (Dutasteride) | 1.233 | 0.909 | 1.674 | 95.2 | 0.0898 | < 0.001 |
| Unger et al., 2016  (Finasteride) | 1.344 | 0.953 | 1.897 | 95.47 | 0.1167 | < 0.001 |

**eTable 2**: Newcastle-Ottawa score rationales for 5 selected studies

|  | **Criteria/ Author** | Garcia-Argibay et al. (2022) | Yeon et al. (2021) | Welk et al (2017) | Hagberg et al., (2017) | Unger et al., (2016) |
| --- | --- | --- | --- | --- | --- | --- |
| Selection:  (Maximum 4 stars) | (1) Representativeness of the exposed cohort | The study included all men aged 50 to 90 years residing in Sweden during the study period, making the cohort truly representative of the target population. | The study used nationwide claims data from South Korea, covering approximately 98% of the national population, which ensures high representativeness of the exposed cohort (5ARI users with BPH). | The exposed cohort (men aged ≥66 years receiving 5-ARIs, finasteride or dutasteride) was drawn from linked administrative data in Ontario, Canada, covering 93,197 men from 2003–2013. This population-based sample is highly representative of older men with BPH in Ontario. | The study utilized the United Kingdom's Clinical Practice Research Datalink (CPRD), a large longitudinal population-based electronic medical record database covering approximately 10 million people, representing the general UK population and ensuring high representativeness of the exposed cohort (5-ARI users). | The exposed cohort (men receiving finasteride in the Prostate Cancer Prevention Trial [PCPT]) consisted of 18,000 men aged ≥55 years, enrolled in a multicenter, randomized, double-blind, placebo-controlled trial |
|  | (2) Selection of the non-exposed cohort | The non-exposed cohort (men not receiving 5-ARIs) was drawn from the same population-based register, ensuring the same community and time- period (2005-2018) | The non-exposed cohort (α-blocker users with BPH) was drawn from the same nationwide claims database and population, ensuring comparability in origin. | The non-exposed cohort was drawn from all remaining men in the same province (Ontario) during the study period and matched to the exposed cohort. | The non-exposed cohort (α-blocker users) was drawn from the same CPRD database, ensuring both cohorts originated from the same community. | The non-exposed cohort was the placebo arm of the same randomized controlled trial (PCPT), drawn from the identical source population as the exposed cohort. |
|  | (3) Ascertainment of exposure | Exposure to 5-ARIs was confirmed via the National Prescription Register (PDR), which records all dispensed medications in Sweden, ensuring secure and reliable records. | Exposure was ascertained through prescription information from nationwide claims data | Exposure (prescription for 5α-reductase inhibitors) was ascertained from the Ontario Drug Benefit Database, which is described as having high accuracy (>99%) and constitutes a secure record. | Exposure was ascertained from recorded prescription details within the CPRD, a comprehensive electronic medical record database known for high accuracy. | Exposure was assigned via randomization in a double-blind, placebo-controlled trial, which is the most secure method for ascertaining exposure. |
|  | (4) Demonstration that outcome of interest was not present at start of study | Included in sensitivity analysis with 4-month lag  Lack of sufficient long lag time to exclude prevalent case | The study explicitly excluded patients with a history of mental disorders (including specific ICD-10 codes for depression) or who were prescribed related medications in the two years prior to the index date, confirming the absence of the outcome at baseline. | The study explicitly excluded patients with a history of depression in the 5 years prior to the index date for the depression analysis, and patients who used 5-ARI two years prior to the index date. | The study explicitly excluded men with a diagnosis of depression, suicidal behaviors, or who received antidepressant prescriptions prior to cohort entry (index date), ensuring the outcome was not present at baseline for eligible participants. Look back period is 1 year | The authors explicitly state a limitation: "Because we were unable to determine baseline conditions, we could not assess whether finasteride should be avoided for certain groups, such as those with baseline depression." They used Medicare claims for follow-up, which were not available at trial registration to rule out pre-existing depression). |
| Comparability  (Maximum 2 stars) | (5) Comparability of cohorts on the basis of the design or analysis | The study adjusted for multiple confounders (e.g., hypertension, obesity, type 2 diabetes, lipid disorders, and year of follow-up start) using time-varying Cox proportional hazards regression models and also propensity score matching | The study utilized 1:1 propensity score (PS) matching to balance a wide range of baseline characteristics, including demographics, healthcare utilization patterns, and numerous comorbidities (e.g., myocardial infarction, hypertension, diabetes, liver disease, neurological disorder, etc.) and concomitant medications known to be potential confounders for depression. The authors reported successful balancing of these covariates after PS matching. | The study used propensity score matching (44/96 covariates, including medical/psychiatric comorbidities, medication use, and healthcare utilization) to control for confounding, with standardized differences <10% post-matching. | The study employed a nested case-control analysis, matching cases and controls on age (year of birth), general practice attended, and index date. Additionally, the analysis adjusted for a comprehensive list of potential confounders, including BMI, smoking status, duration of BPH, various comorbidities (e.g., hypertension, diabetes, CVD, other psychiatric illnesses), and concomitant medications. (Case and control are followed up from CPRD records) | The randomized design inherently balanced most confounders, and baseline characteristics showed no significant differences. |
| Outcome  (Maximum 3 stars) | (6) Assessment of outcome | Previous research showed a 55% sensitivity and 98% specificity for Swedish registries. Only using 1 mode (ICD) to identify outcomes:  depression were defined as the first recorded primary diagnosis through inpatient or outpatient specialist care | Depression as an outcome was ascertained using ICD-10 codes from claims data (F32-34, F38, F412, F432) and/or antidepressant prescriptions (using both drugs and ICD outcomes to enhance validity) | Outcomes (suicide, self-harm, depression) were ascertained through linkage to multiple administrative databases (e.g., Ontario Registrar General-Death database, Canadian Institute for Health Information Discharge Abstract Database) | The outcome, "antidepressant-treated depression," was defined by Read codes for depression diagnosis and receipt of an antidepressant prescription within 90 days (patients must be diagnosed and treated with depression to be qualified as a case) | “An event was identified as any hospital claim—or two or more physician or outpatient claims at least 30 days apart.” |
|  | (7) Was follow-up long enough for outcomes to occur | Majority of patients (64.9%) were followed for at least 48 months | Follow-up extended from the index date (July 2013–June 2015) to June 30, 2017, providing up to 4 years of observation, sufficient for outcomes like depression to manifest in men with BPH. However, median follow-up was not reported | The median follow-up period was approximately 1.6 years, with an interquartile range extending up to 3.5 years, which is sufficient for the observation of psychiatric outcomes. | The study followed patients from 1992 to 2013, with a mean follow-up of greater than 4 years. | The study had a median follow-up of 16 years from trial registration, which is an exceptionally long and sufficient period for depression to be diagnosed and recorded. |
|  | (8) Adequacy of follow up of cohorts | The study used national registers with no reported loss to follow-up, as data linkage ensured complete follow-up until diagnosis, death, emigration, or study end. | The HIRA database ensured comprehensive tracking with no reported loss to follow-up until outcome, death, or study end. However, potential inclusion of patients using 5ARIs for alopecia (not BPH) and limitations in data size per HIRA guidelines introduce potential misclassification | The study used comprehensive administrative data with no reported loss to follow-up, as linkage ensured tracking until outcome, death, emigration, or study end. However, potential misclassification of outcomes (e.g., underreporting of self-harm or depression in administrative data) was not explicitly addressed. Nevertheless, we would expect this bias does not change the results significantly) | The study used a primary care database (CPRD) known for high accuracy and completeness of data, where information is recorded prospectively. Patients were followed until the end of their record, death, or end of the study period. While no specific percentage of loss to follow-up is given, the nature of the continuous and comprehensive data collection within the UK's universal healthcare system suggests minimal uncaptured losses. | The study successfully linked 13 935 (73.8%) of the original trial participants to Medicare claims |
|  | Total score | 7/9 | 9/9 | 9/9 | 9/9 | 8/9 |

**eMethod**

Search term and search strategies:

*Database:* Medline, Embase, Scopus

*Search terms:*

(‘5-alpha reductase inhibitors’ OR ‘finasteride’ OR ‘dutasteride’ OR ‘5-ARIs’) AND (‘depression’ OR ‘depressive symptoms’ OR ‘major depressive disorder’ OR ‘anxiety’ OR ‘suicidal idea’ OR ‘suicide’ OR ‘dementia’ OR ‘Alzheimer disease’ OR ‘vascular dementia’)

*Inclusion and exclusion criteria:*

P: males

I: 5-alpha reductase inhibitors

C: non-5ARI users

O: risk of depression and/or suicide (OR/RR/IRR/HR and 95% CI)

S: case-control, cohort

Search term and search strategies:

Database: Medline, Embase, Scopus

Search terms:

(‘5-alpha reductase inhibitors’ OR ‘finasteride’ OR ‘dutasteride’ OR ‘5-ARIs’) AND (‘depression’ OR ‘depressive symptoms’ OR ‘major depressive disorder’ OR ‘anxiety’ OR ‘suicidal idea’ OR ‘suicide’ OR ‘dementia’ OR ‘Alzheimer disease’ OR ‘vascular dementia’)

Inclusion and exclusion criteria:

P: males

I: 5-alpha reductase inhibitors

C: non-5ARI users

O: risk of depression and/or suicide (OR/RR/IRR/HR and 95% CI)

S: case-control, cohort
